# Using Automated-Machine Learning to Predict COVID-19 Patient Survival: Identify Influential Biomarkers

**DOI:** 10.1101/2020.10.12.20211086

**Authors:** Kenji Ikemura, D.Y. Goldstein, James Szymanski, Eran Bellin, Lindsay Stahl, Yukako Yagi, Mahmoud Saada, Katelyn Simone, Morayma Gil Reyes

## Abstract

**Background:** In a pandemic, it is important for clinicians to stratify patients and decide who receives limited medical resources. In this study, we used automated machine learning (autoML) to develop and compare between multiple machine learning (ML) models that predict the chance of patient survival from COVID-19 infection and identified the best-performing model. In addition, we investigated which biomarkers are the most influential in generating an accurate model. We believe an ML model such as this could be a useful tool for clinicians stratifying hospitalized SARS-CoV-2 patients.

**Methods:** The data was retrospectively collected from Clinical Looking Glass (CLG) on all patients testing positive for COVID-19 through a nasopharyngeal specimen by real-time RT-PCR and admitted between 3/1/2020-7/3/2020 (4376 patients) at our institution. We collected 47 biomarkers from each patient within 36 hours before or after the index time: RT-PCR positivity, and tracked whether a patient survived or not for one month following this time. We utilized the autoML from H2O.ai, an open source package for R language. The autoML generated 20 ML models and ranked them by area under the precision-recall curve (AUCPR) on the test set. We selected the best model (model_var_47) and chose a threshold probability that maximized F2 score to make a binary classifier: dead or alive. Subsequently, we ranked the relative importance of variables that generated model_var_47 and chose the 10 most influential variables. Next, we reran the autoML with these 10 variables and likewise selected the model with the best AUCPR on the test set (model_var_10). Again, threshold probability that maximized F2 score for model_var_10 was chosen to make a binary classifier. We calculated and compared the sensitivity, specificity, and positive predicate value (PPV) for model_var_10 and model_var_47.

**Results:** The best model that autoML generated using all 47 variables was the stacked ensemble model of all models (AUCPR = 0.836). The most influential variables were: systolic and diastolic blood pressure, age, respiratory rate, pulse oximetry, blood urea nitrogen, lactate dehydrogenase, d-dimer, troponin, and glucose. When the autoML was retrained with these 10 most important variables, it did not significantly affect the performance (AUCPR= 0.828). For the binary classifiers, sensitivity, specificity, and PPV of model_var_47 was 83.6%, 87.7%, and 69.8% respectively, while for model_var_10 they were 90.9%, 71.1%, and 51.8% respectively.

**Conclusions:** By using autoML, we developed high-performing models that predict patient mortality from COVID-19 infection. In addition, we identified the most important biomarkers correlated with mortality. This ML model can be used as a decision supporting tool for medical practitioners to efficiently triage COVID-19 infected patients. From our literature review, this will be the largest COVID-19 patient cohort to train ML models and the first to utilize autoML. The COVID-19 survival calculator based on this study can be found at https://www.tsubomitech.com/.

## Introduction

On January 30^th^, 2020, the WHO declared a COVID-19 outbreak which began in Wuhan, China. As of August 1st, 2020, the CDC reported more than 4.5 million cases and 150,000 deaths from COVID-19 in the United States alone [1]. New York City (NYC) became the epicenter in the U.S. with the highest case number and deaths per capita [2]. At our institution, between 3/1/2020-7/3/2020, we admitted 4375 patients who tested positive for COVID-19 and 1088 patients died within 30 days of infection (Figure 1). Many regions worldwide are still fighting the first wave of the pandemic, while other areas that reopened are seeing a resurgence of new cases. In such an emergent situation, it is important for clinicians to triage patients effectively to maximize limited medical resources. In this study, we aimed to find the most important prognostic biomarkers and develop a COVID-19 mortality risk assessment tool using automated machine learning (autoML).

**Figure 1:**
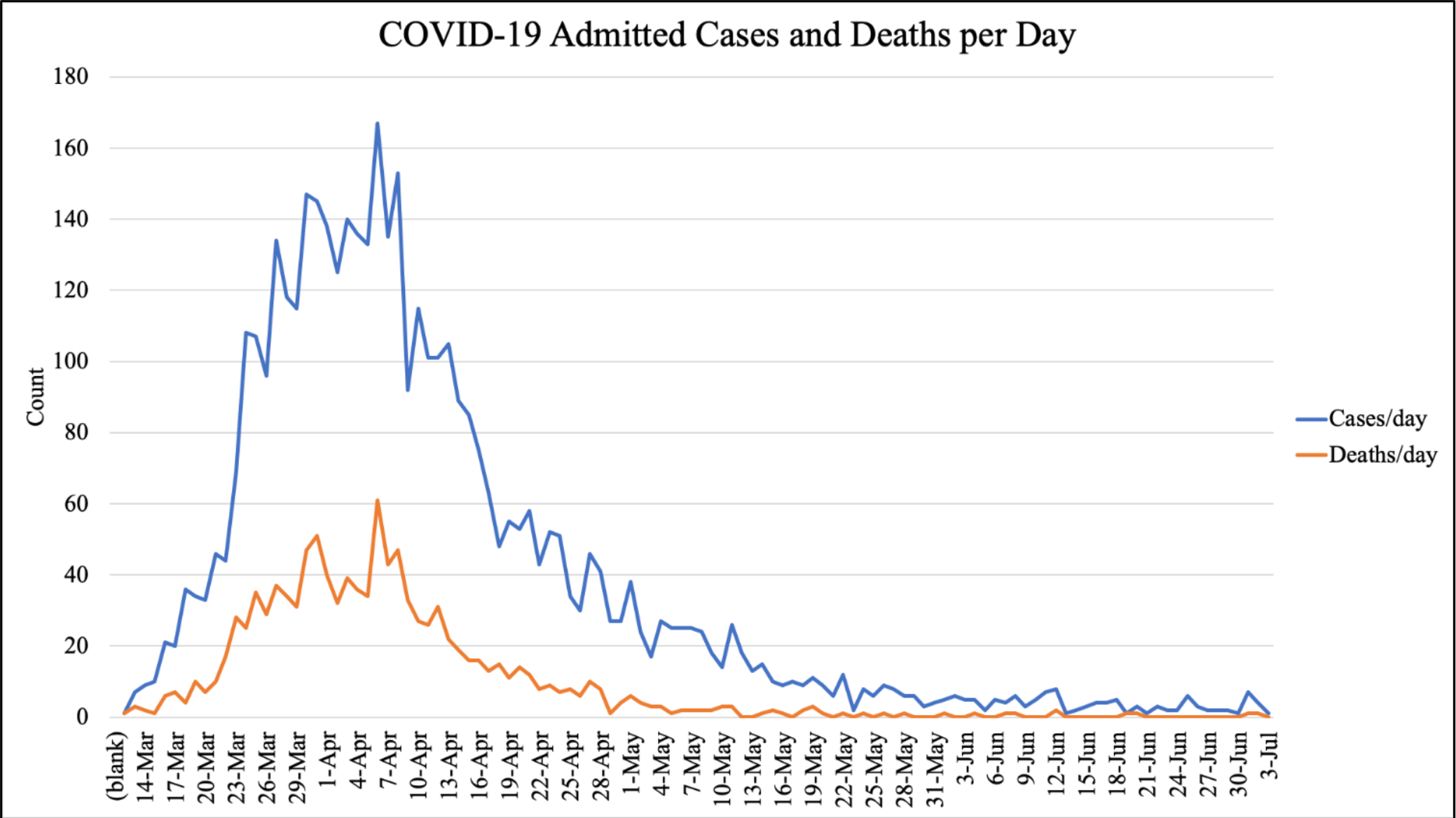
Patients admitted due to COVID-19 infection (blue line) and patient death (red line) per day from March 1^st^ to July 3^rd^ at our institution.

Some of the most frequently reported prognostic indicators in COVID-19 patients include gender, C-reactive protein (CRP), lactic dehydrogenase (LDH), and lymphocyte count. Other inflammatory markers also often appear to be elevated, such as ferritin, inteleukin-6, tumour necrosis factor-alpha, and interferon gamma [3-5]. For example, Zhou et al were one of the first groups that showed many correlations with specific biomarkers that are predictive of morbidity/mortality in 191 patients from Wuhan. They found that older patients with d-dimer levels greater than 1ug/mL, and a higher SOFA (Sequential Organ Failure Assessment) score were associated with greater odds of in-hospital death [6]. However, many papers that have investigated biomarkers are largely focused on Chinese hospitals due to the disease’s initial emergence in Wuhan [7-10]. It is essential to understand the clinical characteristics for more recent and diverse cases considering that the virus strain may have mutated since its first appearance [11].

In recent months, several ML models have been proposed to predict COVID-19 mortality as well as disease severity. In many such ML studies, researchers are only training on one kind of ML model (XGBoost, Random Forest, etc.) and on relatively small datasets with cohorts mainly from China. In addition, many studies evaluated their model performance only on area under the curve (AUC) and without area under the precision recall curve (AUCPR) [3, 12, 13]. AUC should be used if the outcome class (dead or alive) is relatively balanced. If the outcome is skewed, as it is in data showing that most people survive COVID-19 infection, a model should be evaluated with AUCPR.

In our study, we used autoML to generate various machine learning models and automated the hyperparameter tuning and optimization. With autoML, we generated various ML models to compare and choose the best-performing model based on AUCPR. We also used the F2-score to evaluate the binary classifier (dead or alive). Unlike the F1-score, which gives equal weight to positive predictive value (PPV) and sensitivity, the F2-score gives more weight to sensitivity and penalizes the model more for false negatives than false positives. Furthermore, we ranked the variables by importance to understand which variables were the most influential in developing the top-performing models.

While there are many autoML platforms, we chose to use the open source H2O.ai for its diverse ML models in their autoML package [14, 15]. In addition, since the package can be downloaded to a local device, one does not have to upload patient data to the cloud, reducing the risk of exposing it to a third party. The H2O.ai autoML trains and cross-validates on the following ML algorithms: XGBoost, GBM (Gradient Boosting Machine) models, fixed grid of GLMs, a Default Random Forest (DRF), five pre-specified H2O GBMs, near-default Deep Neural Net, an Extremely Randomized Forest (XRT), random grid of XGBoost GBMs, a random grid of H2O GBMs, random grid of Deep Neural Nets, and two Stacked Ensembles - one based on all previously trained models, and another based on the best model of each family [15].

Based on our literature review, this will be the largest COVID-19 patient cohort to train a ML model and the first to utilize autoML.

## Methods

Data collection and analysis was approved by the Albert Einstein College of Medicine Institutional Review Board. The data was collected using Clinical Looking Glass (CLG), an interactive software application developed at Montefiore Medical Center for the evaluation of health care quality, effectiveness, and efficiency. The system integrates clinical and administrative datasets allowing non-statisticians to build temporally sophisticated cohorts and outcomes [16-19].

We queried CLG to find all patients who tested positive for COVID-19 through a nasopharyngeal specimen by real-time RT-PCR and admitted at our institution from 3/1/2020 - 7/3/2020 (4376 patients). The admitted patients tested positive within 24 hours before or after admission. The index time is when RT-PCR resulted positive for COVID-19.

We investigated a total of 47 unique biomarkers after conducting literature reviews, and used the earliest biomarker values available within 36 hours before or after the index time. The outcome of interest was mortality from any cause at one month from index time. We obtained the cycle threshold values (Ct-values) from positive nasopharyngeal specimens collected in accordance with CDC guidance. Real-time RT-PCR was performed using the Hologic Panther Fusion SARS-CoV-2 assay. The variable NLratio is the ratio between neutrophil and lymphocyte count.

We used the open source autoML package from H2O.ai for R programming language [14, 15, 20]. For the purpose of reproducibility, we excluded deep learning method in this study (discussed more in the Discussion section). The autoML is trained on a randomly selected 80% of the dataset (3517 patients, training set) with 10-fold cross-validation. We assigned the autoML to generate 20 machine learning models and rank them in order of performance by AUCPR on the remaining 20% of the dataset (859 patients, test set). As mentioned above, we evaluated the models with AUCPR because there are more patients in our cohort who survived COVID-19 versus those who died. For convenience, we named this best model, generated with 47 variables, “model_var_47.”

To make a binary classifier—dead or alive within 30 days—we chose a threshold probability that maximizes F2 score of the model_var_47. Sensitivity, specificity and PPV were calculated for the binary classifier. As mentioned earlier, F2-score was chosen because, unlike the F1 score which gives equal weight to precision and sensitivity (or recall), the F2-score gives more weight to sensitivity (penalizing the model more for false negatives then false positives).

In addition, we generated variable importance for the top models and chose the ten variables that had the greatest influence in making effective models. Variable importance was determined by calculating the relative influence of each variable: whether that variable was selected to split on during the tree building process, and how much the squared error (over all trees) improved (decreased) as a result [15].

With the 10 chosen variables, we retained the autoML. Again, we assigned the autoML to generate 20 machine learning models and rank them in order of AUCPR on the test set. For convenience, we named the best model, generated with 10 variables, “model_var_10.” Next, to make a binary classifier, we chose a threshold probability that maximizes the F2 score of the model_var_10. Sensitivity, specificity, and PPV were calculated for the binary classifier. Comparison was made between model_var_45 and model_var10.

Regarding how autoML handles missing values for each model is explained in the H20.ai documentation [15].

The workflow of our method is depicted in figure 2.

**Figure 2:**
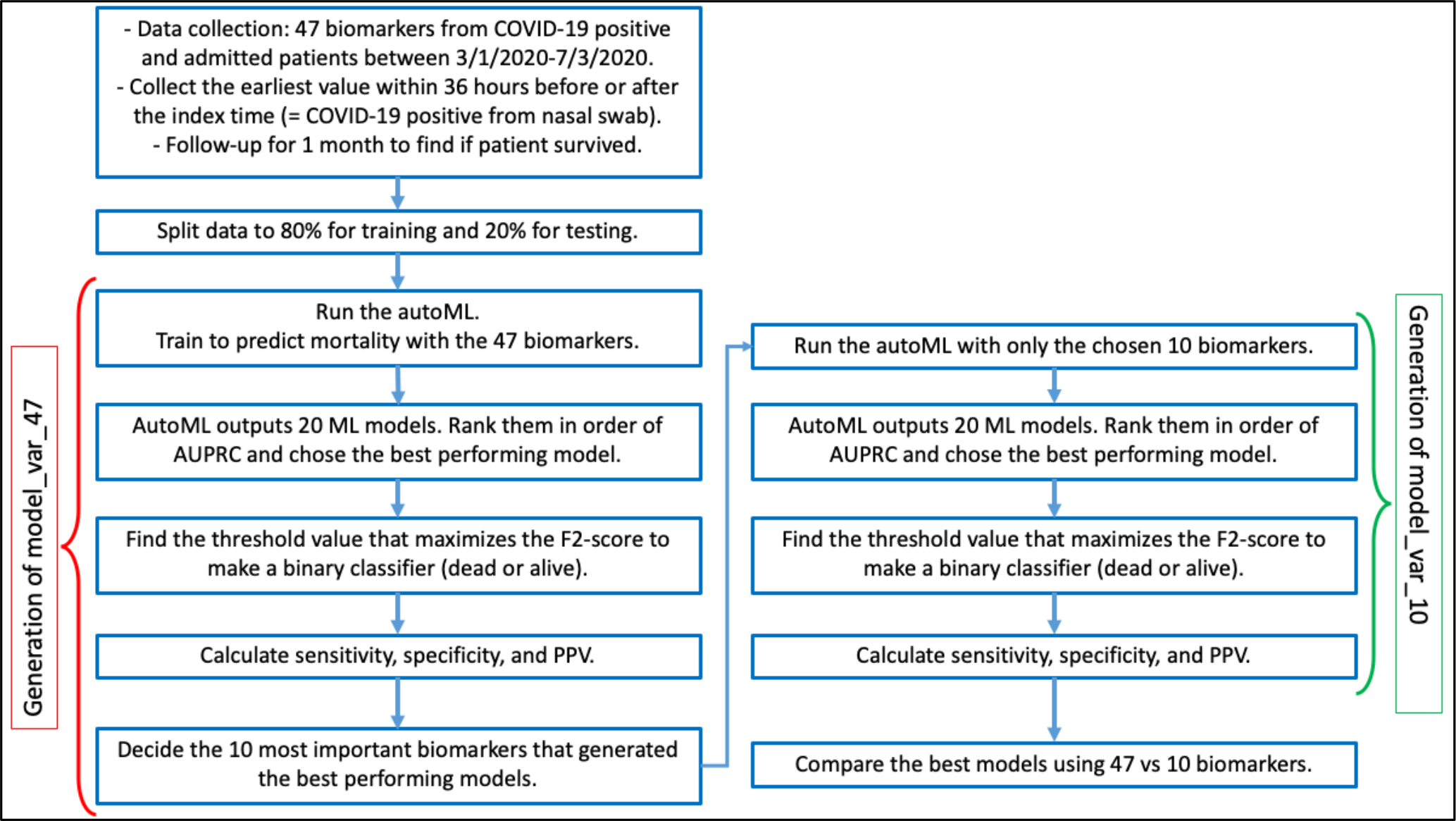
Flowchart summary of our method section.

## Results

Data summary of cases, survival, and biomarkers are presented in Table 1.

**Table 1:** Summary of variables from the cohort. NLratio is the ratio of neutrophil and lymphocyte. The ct_value is the cycle threshold from Hologic Panther Fusion SARS-CoV-2 assay. rr: respiratory rate, mpv: mean platelet volume, NLratio: neutrophil-lymphocyte ratio.

| variable no. | variables | category | n | % missing | mean | sd | median |
| --- | --- | --- | --- | --- | --- | --- | --- |
| 1 | gender |  | 4375 |  |  |  |  |
|  |  | Male | 2326 | 0.00 |  |  |  |
|  |  | Female | 2049 | 0.00 |  |  |  |
| 2 | age |  | 4375 | 0.00 | 63.20 | 17.90 | 65.00 |
| 3 | albumin |  | 4060 | 7.20 | 3.73 | 0.55 | 3.80 |
| 4 | systolicBP |  | 4374 | 0.02 | 120.00 | 24.70 | 122.00 |
| 5 | diastolicBP |  | 4374 | 0.02 | 67.50 | 16.60 | 69.00 |
| 6 | cr |  | 4189 | 4.25 | 2.02 | 2.58 | 1.10 |
| 7 | ddimer |  | 2769 | 36.70 | 3.97 | 5.52 | 1.71 |
| 8 | egfr |  | 4173 | 4.62 | 63.80 | 35.90 | 64.00 |
| 9 | eosinophil |  | 4321 | 1.23 | 0.04 | 0.10 | 0.00 |
| 10 | ferritin |  | 2425 | 44.60 | 1354.00 | 2948.00 | 728.00 |
| 11 | fibrinogen |  | 2009 | 54.10 | 620.00 | 202.00 | 615.00 |
| 12 | hgb |  | 4321 | 1.23 | 12.60 | 2.32 | 12.80 |
| 13 | inr |  | 3805 | 13.00 | 1.23 | 0.95 | 1.10 |
| 14 | lymphocyte |  | 4321 | 1.23 | 1.36 | 4.95 | 1.00 |
| 15 | neutrophil |  | 4321 | 1.23 | 6.75 | 4.20 | 5.70 |
| 16 | NLratio |  | 4321 | 1.23 | 7.84 | 8.74 | 5.67 |
| 17 | platelet |  | 4321 | 1.23 | 235.00 | 109.00 | 215.00 |
| 18 | protein |  | 4045 | 7.54 | 7.07 | 0.78 | 7.10 |
| 19 | pulse |  | 4341 | 0.78 | 99.40 | 21.20 | 99.00 |
| 20 | pulseOx |  | 4339 | 0.82 | 92.80 | 8.33 | 95.00 |
| 21 | rr |  | 4341 | 0.78 | 21.30 | 6.11 | 20.00 |
| 22 | temperature |  | 4338 | 0.85 | 99.20 | 1.69 | 98.80 |
| 23 | wbc |  | 4321 | 1.23 | 8.87 | 7.24 | 7.60 |
| 24 | alt |  | 4055 | 7.31 | 44.80 | 116.00 | 27.00 |
| 25 | ast |  | 4145 | 5.26 | 0.30 | 0.56 | 0.20 |
| 26 | bun |  | 4189 | 4.25 | 32.50 | 32.50 | 20.00 |
| 27 | calcium |  | 4296 | 1.81 | 8.84 | 0.76 | 8.80 |
| 28 | chloride |  | 4145 | 5.26 | 0.30 | 0.56 | 0.20 |
| 29 | crp |  | 4145 | 5.26 | 0.30 | 0.56 | 0.20 |
| 30 | interleukin6 |  | 4145 | 5.26 | 0.30 | 0.56 | 0.20 |
| 31 | ldh |  | 3230 | 26.20 | 455.00 | 358.00 | 381.00 |
| 32 | mcv |  | 4321 | 1.23 | 89.30 | 7.37 | 89.50 |
| 33 | monocyte |  | 4321 | 1.23 | 0.62 | 1.10 | 0.50 |
| 34 | mpv |  | 4217 | 3.61 | 11.00 | 1.11 | 10.90 |
| 35 | procalcitonin |  | 2128 | 51.40 | 2.40 | 7.68 | 0.20 |
| 36 | rdw |  | 4319 | 1.28 | 14.50 | 2.15 | 14.00 |
| 37 | troponin |  | 3625 | 17.14 | 0.06 | 0.26 | 0.01 |
| 38 | ptt |  | 3392 | 22.50 | 35.40 | 14.60 | 32.80 |
| 39 | bmi |  | 4129 | 5.62 | 30.30 | 48.00 | 28.40 |
| 40 | glucose |  | 3736 | 14.60 | 188.00 | 134.00 | 140.00 |
| 41 | direct_bili |  | 4145 | 5.26 | 0.30 | 0.56 | 0.20 |
| 42 | total_bili |  | 4146 | 5.23 | 0.60 | 0.94 | 0.50 |
| 43 | creatinine_kinase |  | 3361 | 23.20 | 640.00 | 3098.00 | 168.00 |
| 44 | pro_bnp |  | 2285 | 47.80 | 2408.00 | 4328.00 | 441.00 |
| 45 | potassium |  | 4209 | 3.79 | 4.43 | 0.74 | 4.30 |
| 46 | charlson_score |  | 4375 | 0.00 | 2.27 | 2.34 | 2.00 |
| 47 | ct_value |  | 1097 | 74.90 | 27.70 | 6.20 | 27.30 |
|  | **survival** |  | **4375** | **0.00** |  |  |  |
|  |  | **dead** | **1088** |  |  |  |  |
|  |  | **alive** | **3287** |  |  |  |  |

### Model_var_47 Performance

The best performing model with 47 variables was Stacked Ensemble of all models with AUCPR = 0.836. This is our model_var_47. This was followed by Stacked Ensemble of best from each ML family (AUCPR = 0.834). After the two Stacked Ensemble models ranked GBM and XGBoost models with AUCPR of 0.830 and 0.825, respectively (Table 2).

**Table 2:** Output of autoML with 47 variables. It is the rank order of models by AUCPR. In addition, it is informing AUC and Logloss.

| rank | model_id | aucpr | auc | logloss |
| --- | --- | --- | --- | --- |
| 1 | StackedEnsemble_AllModels_AutoML_20200816_082220 | 0.836 | 0.919 | 0.300 |
| 2 | StackedEnsemble_BestOfFamily_AutoML_20200816_082220 | 0.834 | 0.918 | 0.302 |
| 3 | GBM_4_AutoML_20200816_082220 | 0.830 | 0.918 | 0.306 |
| 4 | XGBoost_grid__1_AutoML_20200816_082220_model_3 | 0.825 | 0.915 | 0.306 |
| 5 | GBM_1_AutoML_20200816_082220 | 0.820 | 0.912 | 0.317 |
| 6 | XGBoost_grid__1_AutoML_20200816_082220_model_4 | 0.819 | 0.915 | 0.314 |
| 7 | XGBoost_3_AutoML_20200816_082220 | 0.818 | 0.913 | 0.310 |
| 8 | GBM_grid__1_AutoML_20200816_082220_model_3 | 0.817 | 0.911 | 0.317 |
| 9 | XGBoost_grid__1_AutoML_20200816_082220_model_6 | 0.817 | 0.906 | 0.314 |
| 10 | GBM_2_AutoML_20200816_082220 | 0.817 | 0.916 | 0.306 |
| 11 | XGBoost_grid__1_AutoML_20200816_082220_model_2 | 0.813 | 0.906 | 0.347 |
| 12 | XGBoost_grid__1_AutoML_20200816_082220_model_1 | 0.813 | 0.912 | 0.314 |
| 13 | GBM_5_AutoML_20200816_082220 | 0.812 | 0.914 | 0.317 |
| 14 | XGBoost_grid__1_AutoML_20200816_082220_model_5 | 0.809 | 0.909 | 0.319 |
| 15 | XRT_1_AutoML_20200816_082220 | 0.807 | 0.902 | 0.343 |
| 16 | GBM_3_AutoML_20200816_082220 | 0.807 | 0.910 | 0.319 |
| 17 | GBM_grid__1_AutoML_20200816_082220_model_2 | 0.803 | 0.900 | 0.338 |
| 18 | GBM_grid__1_AutoML_20200816_082220_model_1 | 0.796 | 0.902 | 0.326 |
| 19 | DRF_1_AutoML_20200816_082220 | 0.796 | 0.892 | 0.351 |
| 20 | XGBoost_2_AutoML_20200816_082220 | 0.788 | 0.903 | 0.328 |

Max F2-score of our model_var_47 was 0.804 with the threshold probability of 0.215. The binary classifier with this threshold had sensitivity, specificity, and PPV of 83.6%, 87.7%, and 69.8%, respectively (Table 3).

**Table 3:**
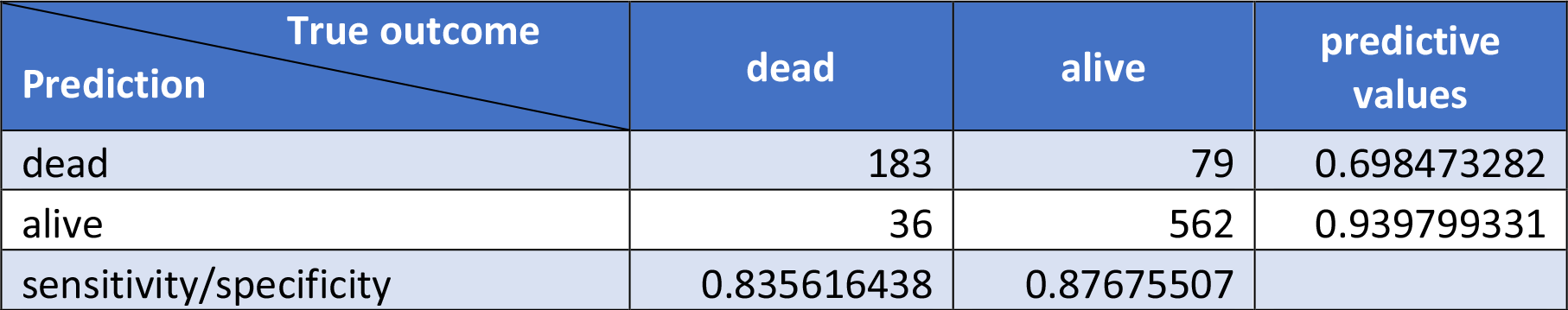
Confusion matrix of model_var_47. To make a binary classifier, we chose a threshold of 0.215 that maximized F2 score of 0.804. Sensitivity = 83.6%, specificity = 87.7%, positive predictive value = 69.8%, negative predictive value = 94.0%.

### Variable Importance Ranking

Ensemble Models cannot generate a variable importance ranking. However, we can generate the variable importance ranking for the XGBoost and GBM models (Table 4). For both models, age and vital signs (blood pressure, pulse Ox, and respiratory rate (RR)) ranked high. In addition, biomarkers such as BUN, LDH, D-dimer, creatinine (Cr), EGFR, troponin, pro-BNP, glucose, and procalcitonin also appeared high in the rank for many models. To have at least one biomarker for cardiac and renal function, we chose troponin and BUN to be included in the 10 variables. Glucose was also chosen for its high rank and ease of measuring in clinical setting. Therefore, our chosen 10 variables were: systolic and diastolic blood pressure, age, pulse Ox, respiratory rate, LDH, BUN, D-dimer, troponin, and glucose.

**Table 4:**
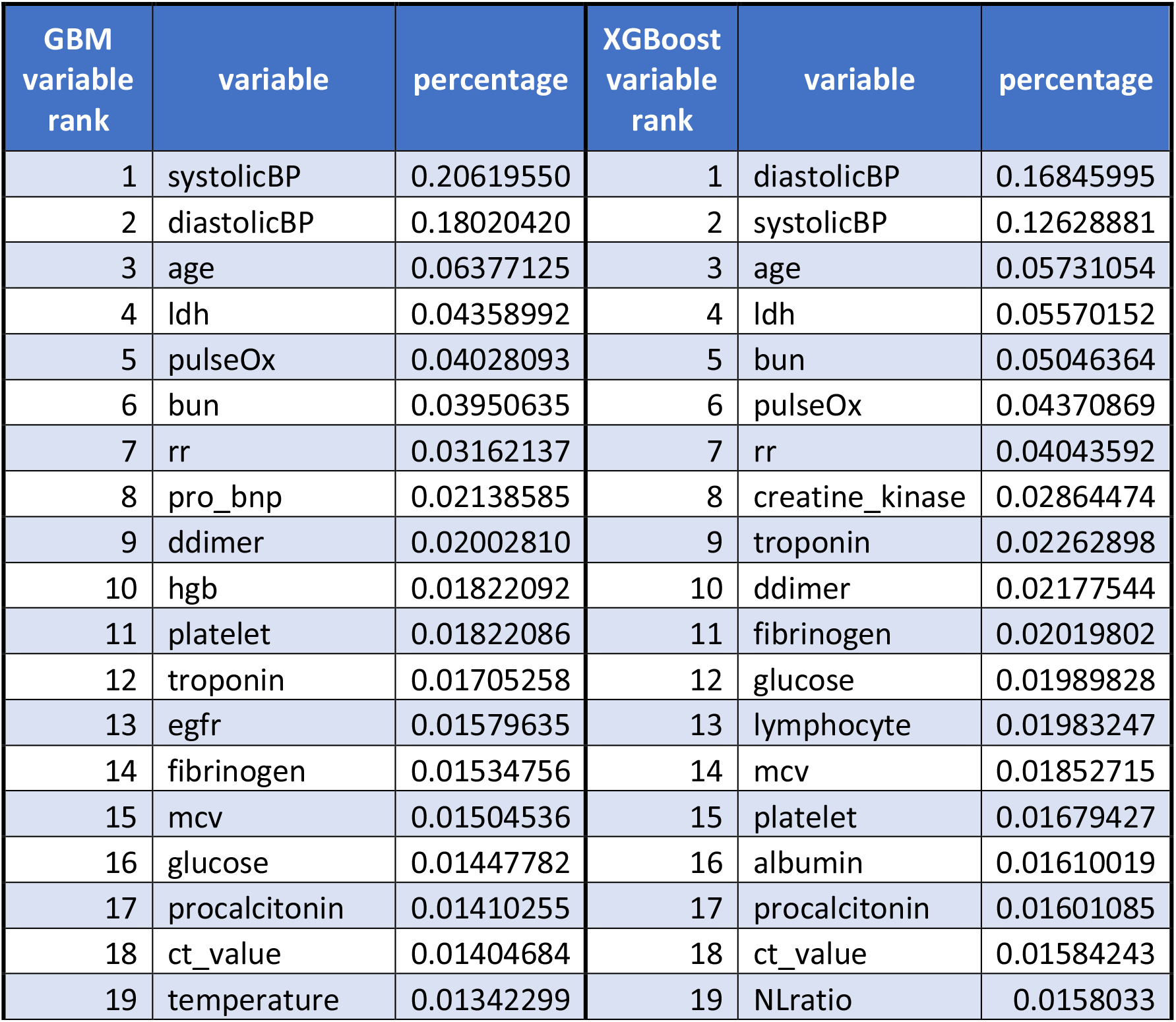
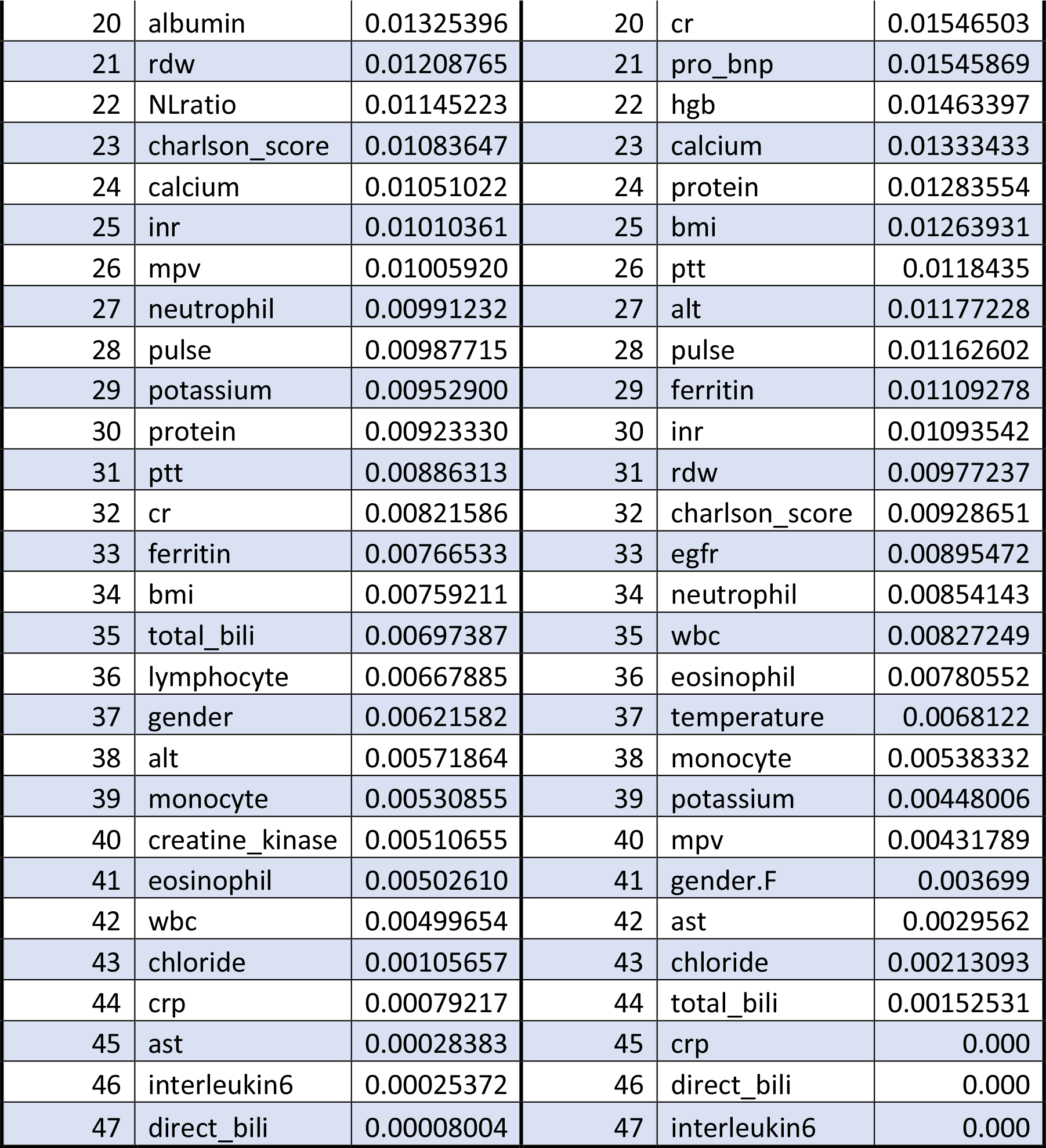
Side by side comparison of variable importance ranking between best performed GBM and XGBoost model. rr: respiratory rate, mpv: mean platelet volume, NLratio: neutrophil-lymphocyte ratio.

### Model_var_10 Performance

Next, we reran the autoML with the chosen 10 variables and ranked the models in order of AUCPR. The GBM model performed the best with AUCPR = 0.828. This is our model_var_10. Table 5 shows the output from autoML showing the rank order of models by AUCPR.

**Table 5:** Output of autoML with ran with 10 variables. It is showing the rank order of models by AUCPR. In addition, it is informing AUC and Logloss.

| rank | model_id | aucpr | auc | logloss |
| --- | --- | --- | --- | --- |
| 1 | GBM_2_AutoML_20200816_084209 | 0.828 | 0.911 | 0.315 |
| 2 | GBM_3_AutoML_20200816_084209 | 0.818 | 0.908 | 0.318 |
| 3 | StackedEnsemble_BestOfFamily_AutoML_20200816_084209 | 0.818 | 0.915 | 0.313 |
| 4 | StackedEnsemble_AllModels_AutoML_20200816_084209 | 0.818 | 0.916 | 0.311 |
| 5 | GBM_grid__1_AutoML_20200816_084209_model_3 | 0.817 | 0.912 | 0.313 |
| 6 | XGBoost_grid__1_AutoML_20200816_084209_model_1 | 0.816 | 0.912 | 0.311 |
| 7 | GBM_grid__1_AutoML_20200816_084209_model_2 | 0.814 | 0.906 | 0.318 |
| 8 | XGBoost_grid__1_AutoML_20200816_084209_model_5 | 0.810 | 0.910 | 0.321 |
| 9 | XGBoost_grid__1_AutoML_20200816_084209_model_6 | 0.809 | 0.909 | 0.316 |
| 10 | XGBoost_grid__1_AutoML_20200816_084209_model_4 | 0.809 | 0.907 | 0.319 |
| 11 | XGBoost_3_AutoML_20200816_084209 | 0.808 | 0.908 | 0.317 |
| 12 | GBM_4_AutoML_20200816_084209 | 0.807 | 0.903 | 0.323 |
| 13 | GBM_1_AutoML_20200816_084209 | 0.804 | 0.909 | 0.317 |
| 14 | XGBoost_1_AutoML_20200816_084209 | 0.803 | 0.904 | 0.325 |
| 15 | GBM_5_AutoML_20200816_084209 | 0.801 | 0.912 | 0.323 |
| 16 | DRF_1_AutoML_20200816_084209 | 0.798 | 0.904 | 0.359 |
| 17 | XGBoost_grid__1_AutoML_20200816_084209_model_3 | 0.796 | 0.904 | 0.324 |
| 18 | GBM_grid__1_AutoML_20200816_084209_model_1 | 0.792 | 0.902 | 0.330 |
| 19 | XRT_1_AutoML_20200816_084209 | 0.790 | 0.905 | 0.362 |
| 20 | XGBoost_2_AutoML_20200816_084209 | 0.787 | 0.896 | 0.334 |

Max F2-score of our model_var_10 was 0.790 with the threshold probability of 0.151. The binary classifier with this threshold had sensitivity, specificity, and positive predictive value of 90.9%, 71.1%, and 51.8%. (Table 6).

**Table 6:** Confusion matrix of model_var_10. To make a binary classifier, we chose a threshold of 0.151 that maximized F2 score of 0.790. Sensitivity = 90.9%, specificity = 71.1%, positive predictive value = 51.8%, negative predictive value = 95.8%.

| Prediction \ True outcome | dead | alive | predictive values |
| --- | --- | --- | --- |
| dead | 199 | 185 | 0.518229167 |
| alive | 20 | 456 | 0.957983193 |
|  | 0.908675799 | 0.711388456 |  |

Figure 3 shows the comparison of model_var_47 and model_var_10 by AUCPR.

**Figure 3:**
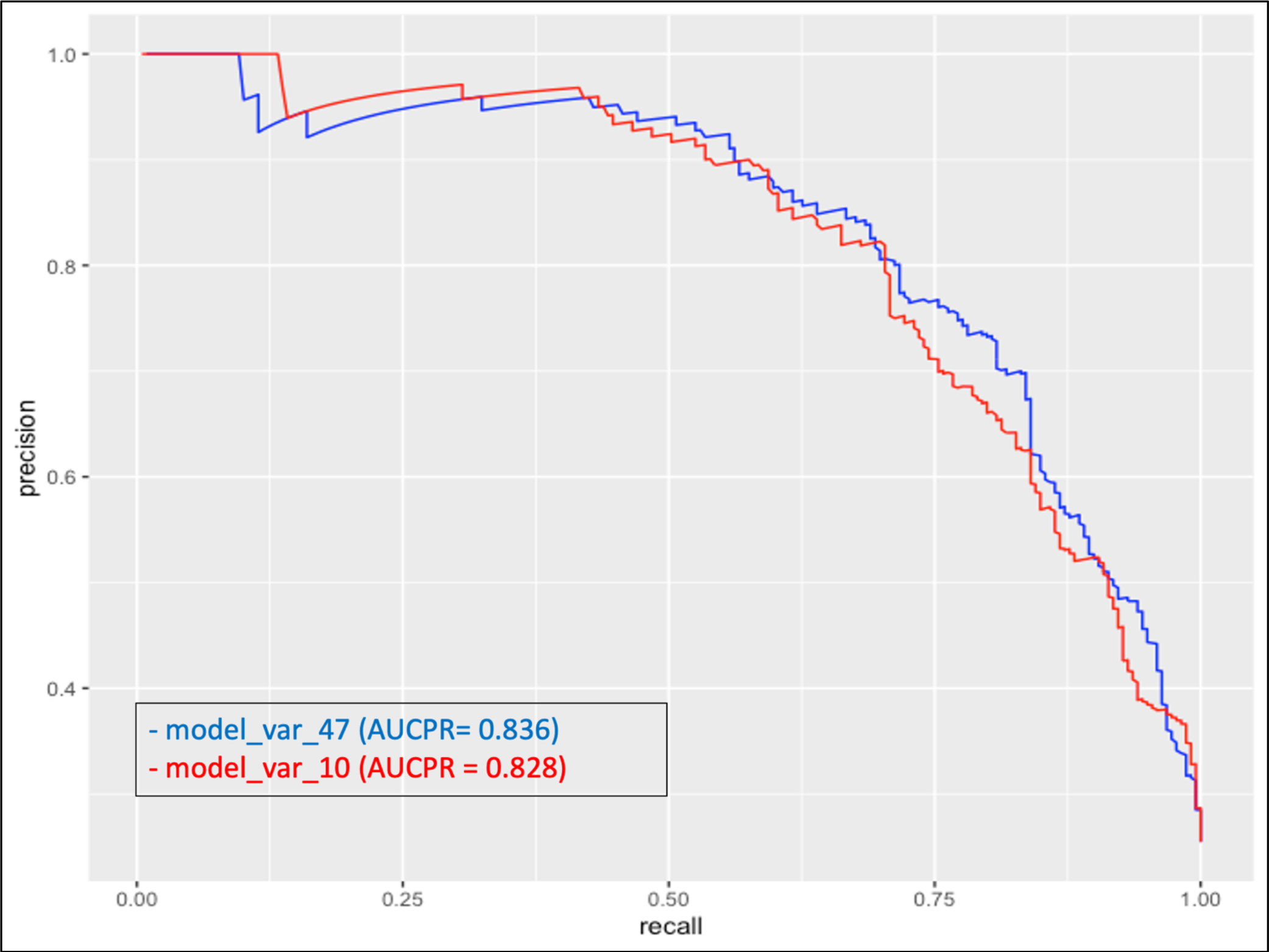
Comparing AUCPR of model_var_47 and model_var_10.

## Discussion

### Principal Results

By using autoML, we were able to generate multiple ML models, automate the optimization of hyperparameter tuning, and compare models by their performance. In addition, we were able to extract the variables that are most predictive of patient mortality.

The model_var_47 and model_var_10 both predict mortality with high performance using clinical measurements collected early within a patient’s hospital admission. When the autoML was retrained with the 10 chosen variables, it did not significantly affect the performance of the model. This suggests that these biomarkers are essential in gauging patient’s severity from the infection. Our models use commonly available laboratory results and do not require imaging results or advanced testing. We believe an early and convenient risk assessment of patient mortality can allow physicians to triage and prioritize resources in a highly congested medical system.

Of the models generated by autoML, Stacked Ensemble performed the best with all 47 variables and GBM model performed the best with the 10 chosen variables. Deep learning was eliminated in this study for the purpose of reproducibility. However, when autoML ran with deep learning, it did not perform as well as GBM or XGBoost models. Deep learning’s performance was at best AUCPR of 0.807 on the test set.

For both GBM and XGBoost, age and vital signs had significant influence in predicting patient mortality. LDH came at the top as the most reliable inflammatory marker. Cardiac (troponin and pro-BNP) and renal (Cr and EGFR) markers were influential, supporting studies from [21] and [22]. D-dimer ranked high in many models, which supports studies that found COVID-19 can promote coagulopathy [23, 24]. Glucose also ranked relatively high in our models supporting the findings that fasting blood glucose, regardless of previous diagnosis of diabetes or not, can be affected from COVID-19 infection [25]. Other variables that often came up within top 15 in importance ranking were, creatine kinase, fibrinogen, hemoglobin, and platelet. Regarding Ct-value, because it is not standardized across RT-PCR platforms, results can not be compared across different assays. In this study, we only used one platform (the Hologic Panther Fusion) and therefore there were more missing values (74% missing). It requires larger dataset to find if magnitude of a Ct-value have clinical implications.

Some variables, especially in the lower rank, seem to be contributing to the model but it is not clear how they are clinically influenced from COVID-19 infection. For example, MCV and calcium level (rank 15 and 24 in GBM model) appear to affect the model, but because ML models are a non-linear combination of variables, it is difficult to understand how they are contributing and if there is clinical significance. Further investigation is needed on how these biomarkers are altered in COVID-19 infected patients. However, it is interesting to note that ML models can identify these alterations.

## Limitations

We recognize there are limitations to our study. Fibrinogen, procalcitonin, and Ct-values were missing in more than 50% of our cohort. H2O.ai has internal imputation method to fill in these missing values [15]. Our cohort was limited to patients severe enough to be admitted, and findings may not generalize to all COVID-19 infected patients. Finally, the nature of training ML models involves randomization. In this study, we presented one representative autoML run. To check for consistency, we tested multiple random splitting between training and test set and retrained the autoML each time. We found that Stacked Ensemble models usually perform the best, followed by GBM and XGBoost. The overall performance of each model was minimally altered from the randomization process. Variable ranking shifted slightly with each autoML run, especially in the lower-ranking variables. However, the 10 most important biomarkers we chose consistently ranked at the top.

## Future Work

We are working towards additional goals to make the model more robust and user friendly for clinicians. First, we are in process of gathering data from other institutions and countries to make our model more generalizable. We imagine that institutions with many COVID-19 patients can develop their own ML models specialized for their population; institutions with minimal COVID-19 cases can use a more generalized model trained from a metadata. Second, we are working to implement reinforcement learning into our model. Reinforcement learning will allow us to update the model in real time as we accumulate more data. This will make the model responsive to a rapidly changing environment. Lastly, we are working to open the black box of ML models to understand how they are making such highly accurate decisions. For example, we would like to know: what is the cut-off value for each variable to be considered important? How are variables related to each other? Deciphering the black box of ML models is a research field of its own. However, it will allow us to use the models more practically, and possibly provide insight into the mechanism of disease of COVID-19 infection [26].

## Conclusion

We generated high-performing ML models that predicts mortality of COVID-19 infected patients using autoML. We also identified important variables that are strongly associated with patient outcomes. This is a proof of concept that autoML is an efficient, effective, and informative method to generate ML models and gain insight into the disease process. A model such as this may help clinicians triage patients in the current pandemic.

## Data Availability

Data is availably for reproducibility.

